# Dosimetric Analysis of Fast Forward Breast Radiotherapy Using 3D-CRT with Deep Inspiration Breath Hold (DIBH)

**DOI:** 10.1101/2025.07.18.25331768

**Authors:** M Ajithkumar, Subhankar Show, Aaditya Prakash

## Abstract

**Introduction:** Fast Forward radiotherapy (26 Gy in 5 fractions) has gained widespread acceptance for early-stage breast cancer due to its convenience and efficacy. However, in left-sided cases, there is a risk of radiation-induced cardiac and pulmonary toxicity due to the proximity of the heart and lungs. The voluntary deep inspiration breath hold (vDIBH) technique increases thoracic volume and heart-chest wall distance, offering a non-invasive method to minimize organ-at-risk exposure.

**Objective:** To evaluate the dosimetric benefits of 3D-conformal radiotherapy (3D-CRT) combined with voluntary deep inspiration breath hold (vDIBH) in minimizing radiation dose to the heart and lungs during ultra-hypofractionated whole breast irradiation in patients with left-sided breast cancer.

**Materials and Methods:** We retrospectively analyzed 28 patients with left-sided breast cancer treated with 3D-CRT under vDIBH, receiving 26 Gy in 5 fractions. Target volumes and organs at risk (OARs) were contoured on vDIBH CT scans. Treatment planning was performed using tangential fields. Dosimetric parameters for the planning target volume (PTV), heart, ipsilateral lung, contralateral breast, and spinal cord were evaluated using dose-volume histograms (DVHs). Reproducibility of vDIBH was ensured by using in-room lasers, skin tattoos, and cine-mode imaging for intra-fraction verification.

**Results:** Mean PTV V95% was 97.74 ± 1.74%, with a mean dose of 26.61 ± 0.25 Gy. The mean heart dose was 3.97 ± 0.82 Gy, with V25% at 13.28 ± 3.44%. Ipsilateral lung Dmean was 8.26 ± 0.76 Gy, and V30% was 32.81 ± 2.85%. Contralateral breast V5% was 5.19 ± 7.06%. The spinal cord received a negligible dose (Dmax 0.49 ± 0.12 Gy). All patients tolerated vDIBH well, with no delays or treatment interruptions.

**Conclusion:** 3D-CRT with vDIBH offers a practical, reproducible, and cost-effective approach for delivering hypofractionated radiotherapy in left-sided breast cancer. It ensures excellent target coverage while significantly reducing radiation dose to critical structures such as the heart and lungs, supporting its use as a standard practice even in resource-constrained settings.

## 1. Introduction

Postoperative radiation therapy (RT) is a cornerstone in the management of early-stage breast cancer, significantly reducing the risk of local recurrence and improving overall survival [14, 16]. In recent years, hypofractionated regimens, such as Fast Forward Breast Radiotherapy (26 Gy in 5 fractions over 1 week), have become increasingly popular due to their convenience, reduced treatment time, and comparable efficacy to conventional schedules. In cases of left-sided breast cancer, the heart and left anterior descending (LAD) artery lie in close proximity to the chest wall, increasing the potential for radiation-related cardiac injury. Several large studies have demonstrated that cardiac doses from breast cancer radiotherapy are generally higher for left-sided than for right-sided tumors, with long-term data indicating that the cumulative risk of cardiac death at 20 years is 6.4% for left-sided cases compared to 3.6% for right-sided counterparts [8].

To mitigate this risk, various strategies have been developed, among which the voluntary Deep Inspiration Breath Hold (vDIBH) technique has emerged as a simple, reproducible, and effective method. vDIBH involves asking the patient to take and hold a deep breath during radiation delivery, which expands the lungs and moves the heart inferiorly and posteriorly, thereby increasing the distance between the heart and the treatment field [2, 3, 7]. This approach effectively limits the amount of radiation reaching the heart and left anterior descending (LAD) artery. McIntosh et al. reported that use of vDIBH in 3D-conformal whole-breast radiotherapy resulted in a 7% reduction in mean heart dose and a 9% reduction in mean LAD dose compared to free breathing [1]. In addition to reducing cardiac dose, vDIBH has also been associated with a decrease in the volume of lung irradiated, thereby minimizing the risk of pulmonary toxicity [2, 3]. The vDIBH technique, in particular, has gained traction in clinical settings because it is simple to implement, requires no specialized external equipment, and is cost-effective. With appropriate patient coaching and immobilization, this technique can achieve reproducible breath hold levels and stable anatomy throughout treatment [3–5]. Despite advances in highly conformal techniques like IMRT, issues such as sensitivity to intrafraction motion, coverage of the flash region, and increased low-dose exposure remain concerns, particularly in breast treatment [10]. In contrast, 3D-CRT combined with vDIBH provides a balanced approach by maintaining adequate target coverage while effectively sparing organs at risk. Furthermore, the field-in-field (FIF) technique, a variation of 3D-CRT that uses additional subfields with manually shaped apertures, improves dose homogeneity, reduces acute skin toxicity, and enhances cosmetic outcomes [16].

The risk of cardiac toxicity from breast radiotherapy is influenced by both the treatment technique and the dose-volume relationship. Additionally, the administration of cardiotoxic systemic agents such as anthracyclines, trastuzumab, and taxanes highlights the critical need to limit cardiac radiation exposure [10, 14]. The vDIBH technique allows for substantial sparing of the heart without compromising target coverage or increasing dose to the contralateral breast [11]. As such, it has been adopted as the standard of care for left-sided breast cancer RT in many institutions. While advanced techniques like IMRT and VMAT offer superior dose conformity, 3D-CRT was chosen for this study due to its robustness against intrafraction motion, simpler quality assurance, and cost-effectiveness. This study, titled “Dosimetric Analysis of Fast Forward Breast Radiotherapy Using 3D-CRT with Deep Inspiration Breath Hold (DIBH),” presents a retrospective dosimetric analysis of patients treated with 3D-CRT using the vDIBH technique, focusing on the extent of cardiac sparing achieved.

## 2. Materials and Methods

### 2.1 Patient Selection

We retrospectively reviewed 28 patients with left-sided breast cancer who underwent mastectomy followed by irradiation to the whole breast using an ultra-hypofractionated regimen (26 Gy in 5 fractions). All patients had early-stage disease, classified as T1–T2, N0, M0 based on the AJCC TNM staging system. Patient ages ranged from 35 to 60 years, with a median age of 47 years. Patients with pre-existing cardiac disease (e.g., coronary artery disease, cardiomyopathy), severe pulmonary comorbidities (COPD GOLD ≥3), or prior thoracic radiotherapy were excluded to isolate the dosimetric impact of vDIBH. Additionally, those unable to tolerate breath-hold durations >20 seconds during training were deemed ineligible. Regional lymph nodes, including the supraclavicular, undissected axillary, and internal mammary (IM) nodes, were not included in the treatment volumes. All relevant target volumes were contoured on vDIBH planning CT scans using the Eclipse Treatment Planning System (Varian Medical Systems, Palo Alto, CA). Treatment planning was performed using 3D-CRT with tangential fields to cover the breast tissue [7].

### 2.2 Patient Positioning and Simulation

All patients were positioned on an inclined breast board with both arms raised above the head and immobilized using dedicated arm supports to ensure reproducibility during simulation and treatment [1]. The borders of the tangential treatment fields were determined clinically by the radiation oncologist and marked with radiopaque wires. Computed tomography (CT) simulation was performed with a slice thickness of 3 mm, encompassing anatomical regions from the mid-neck to the mid-abdomen [14].

### 2.3 Laser Alignment and Breath-Hold Reproducibility

To verify and monitor chest wall excursion during vDIBH, an additional laser was mounted on a tripod to avoid interference from the gantry. A bright ink marker line was drawn on the patient’s skin posterior to the tattooed alignment mark, based on the depth measured during CT simulation. During deep inspiration, proper alignment of the laser with the ink mark confirmed consistent breath-hold positioning. This alignment was observed remotely via a magnification camera by the radiation therapist throughout beam delivery. If misalignment (e.g., indicating expiration) was observed, the beam was manually interrupted to maintain accuracy [2].

### 2.4 Patient Compliance and Training

Prior to CT simulation, all patients were educated about the vDIBH technique, as it requires active patient participation to achieve and maintain consistent breath-holds during each treatment field. Each patient was instructed to complete two regular breaths, followed by a deep inhalation held for 20 seconds during simulation. Planning scans were acquired in both the vDIBH and free-breathing (FB) states to support dosimetric evaluation and treatment design [3, 11].

### 2.5 Treatment Planning and Setup

The initial treatment setup was based on skin markings and alignment with in-room lasers. The variation in inspiratory depth between free breathing (FB) and voluntary deep inspiration breath hold (vDIBH) was assessed and indicated on the patient’s skin surface. During each treatment, the patient was instructed via audiovisual communication from the treatment console to breathe in and hold until the skin mark aligned with the in-room laser. The radiation therapists observed breath hold positioning via a live video feed and instructed the patient accordingly. In addition to routine, an Electronic Portal Imaging Device (EPID) was operated in CINE mode to capture a sequence of images during beam-on time for intra-fraction verification [3].

### 2.6 CT Image Matching and PTV Margin Calculation

The translational setup errors were calculated by comparing CBCT (cone beam CT) images with the planning CT. The planning target volume (PTV) margins were calculated using the Van Herk formula:

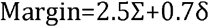

Where Σ represents the systematic error and δ represents the random error [18].

### 2.7 Dosimetric Evaluation

Treatment plans were evaluated using dose volume histograms (DVHs). The target coverage was assessed using the following parameters: PTV: V95%, V105%, V107%, and Dmax, Heart: V5%, V25%, and Mean Dose, Left Lung: V30% and Dmean, Right Breast: V5%. Each patient was treated with a prescribed dose of 26 Gy delivered in 5 fractions using 3D-CRT tangential fields. Dose constraints were evaluated to assess the extent of cardiac and pulmonary sparing achieved with the vDIBH technique [13]

## 3. Results

Dosimetric outcomes were evaluated for 28 patients with left-sided breast cancer treated with 3D-CRT under vDIBH, prescribed to 26 Gy in 5 fractions. Dose volume histogram data were extracted from the treatment planning system.

(Table 1):

V95% - volume of PTV receiving ≥95% of the prescribed dose; V107% - volume of PTV receiving ≥107% of the prescribed dose; Dmean - mean dose; Dmax - maximum dose; V5%, V25%, V30% percentage volume receiving at least 5%, 25%, or 30% of the prescribed dose, respectively. All dose values are expressed in Gy. Values are presented as mean ± standard deviation.

From Table 1, Target Volume Coverage of PTV achieved robust coverage: V95% was 97.74 ± 1.74%, meeting RTOG 1005 protocol requirements[19].Dmean was 26.61 ± 0.253 Gy, aligning with the prescribed dose.V107% was 6.31 ± 5.76%, consistent with acceptable dose inhomogeneity for field in field modulation. Heart Dose Parameters, vDIBH effectively reduced cardiac exposure: V5% was 40.72 ± 12.98%,V25% was 13.28 ± 3.44%.The mean heart dose was 3.98 ± 0.83 Gy, well below the 5.0 Gy constraint for ultra-hypofractionated breast radiotherapy (RTOG 1005).Left Lung Dose Parameters, The ipsilateral left lung showed moderate involvement due to tangential field geometry: V30% was 32.81 ± 2.85%.The mean lung dose was 8.27 ± 0.77 Gy, consistent with expected values for conformal planning. Contralateral Breast Dose,The contralateral breast received minimal scatter dose: V5% was 5.19 ± 7.06%, indicating effective sparing to reduce the risk of secondary malignancy. Spinal Cord Dose, The spinal cord included for completeness, received a negligible Dmax of 0.49 ± 0.13 Gy, well below tolerance limits.

**TABLE 1:** Dosimetric Parameters for Target and Organs at Risk in Patients Treated with 3D-CRT Using vDIBH.

| TREATMENT PLANNING |  | 3DCRT |
| --- | --- | --- |
| PTV | V95% | 97.74±1.74 |
|  | V107% | 6.31±5.76 |
|  | D<sub>mean</sub>(cGy) | 26.61±0.25 |
| HEART | V5% | 40.72±12.98 |
|  | V25% | 13.28±3.44 |
|  | D<sub>mean</sub>(cGy) | 3.97±0.82 |
| LT LUNG | V30% | 32.81±2.85 |
|  | D<sub>mean</sub>(cGy) | 8.26±0.76 |
| RT BREAST | V5% | 5.19±7.06 |
| SPINALCORD | D<sub>max</sub>(cGy) | 0.49±0.12 |

Figures 1, 2, 3 Axial CT images and isodose distribution illustrating the impact of vDIBH in left-sided breast radiotherapy.

**FIGURE 1:**
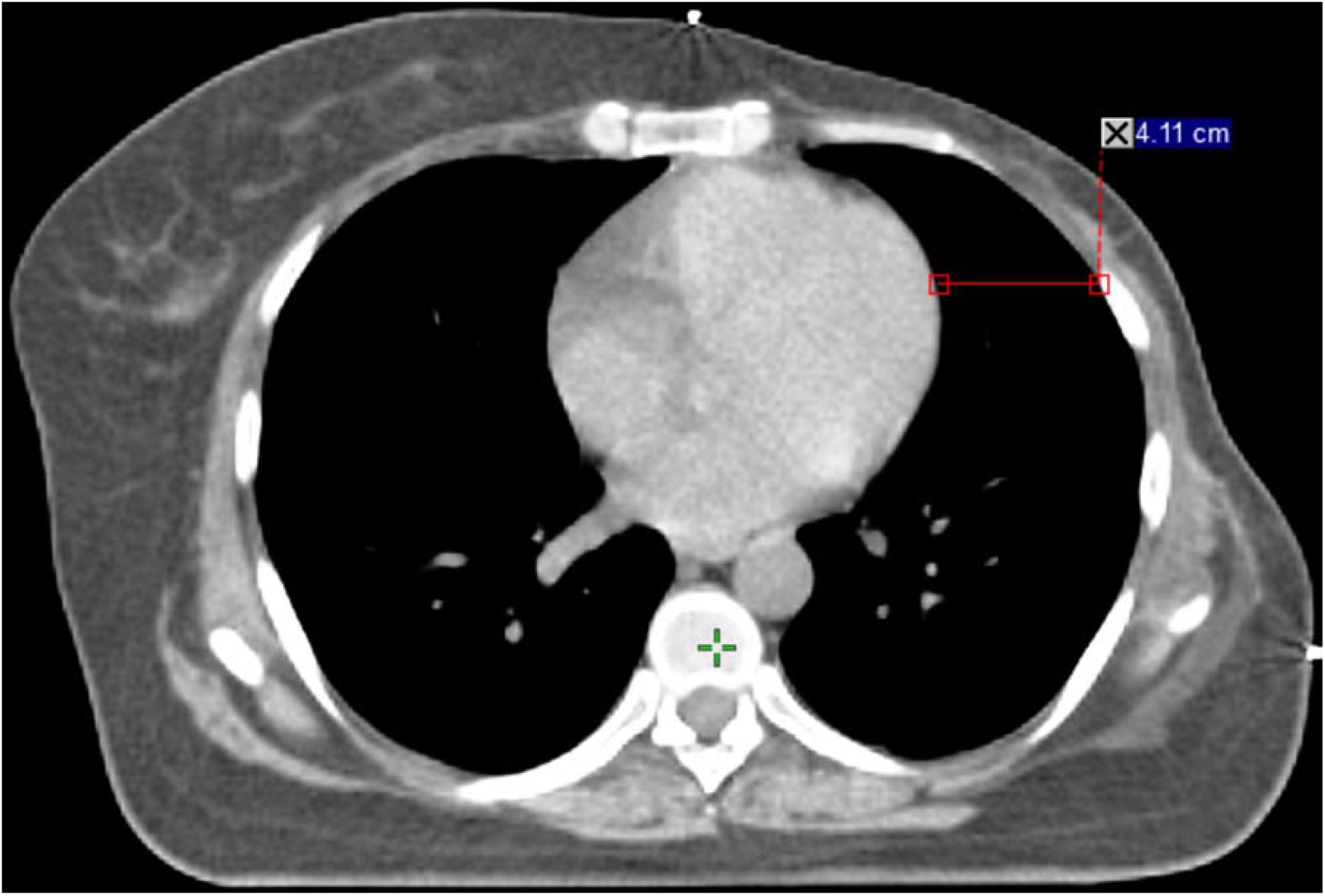
CT image acquired during voluntary deep inspiration breath hold (vDIBH). This axial planning CT shows an increased separation of 4.11 cm between the anterior surface of the heart and the chest wall, demonstrating the effectiveness of vDIBH in displacing the heart posteriorly and inferiorly, away from the tangential radiation fields.

**FIGURE 2:**
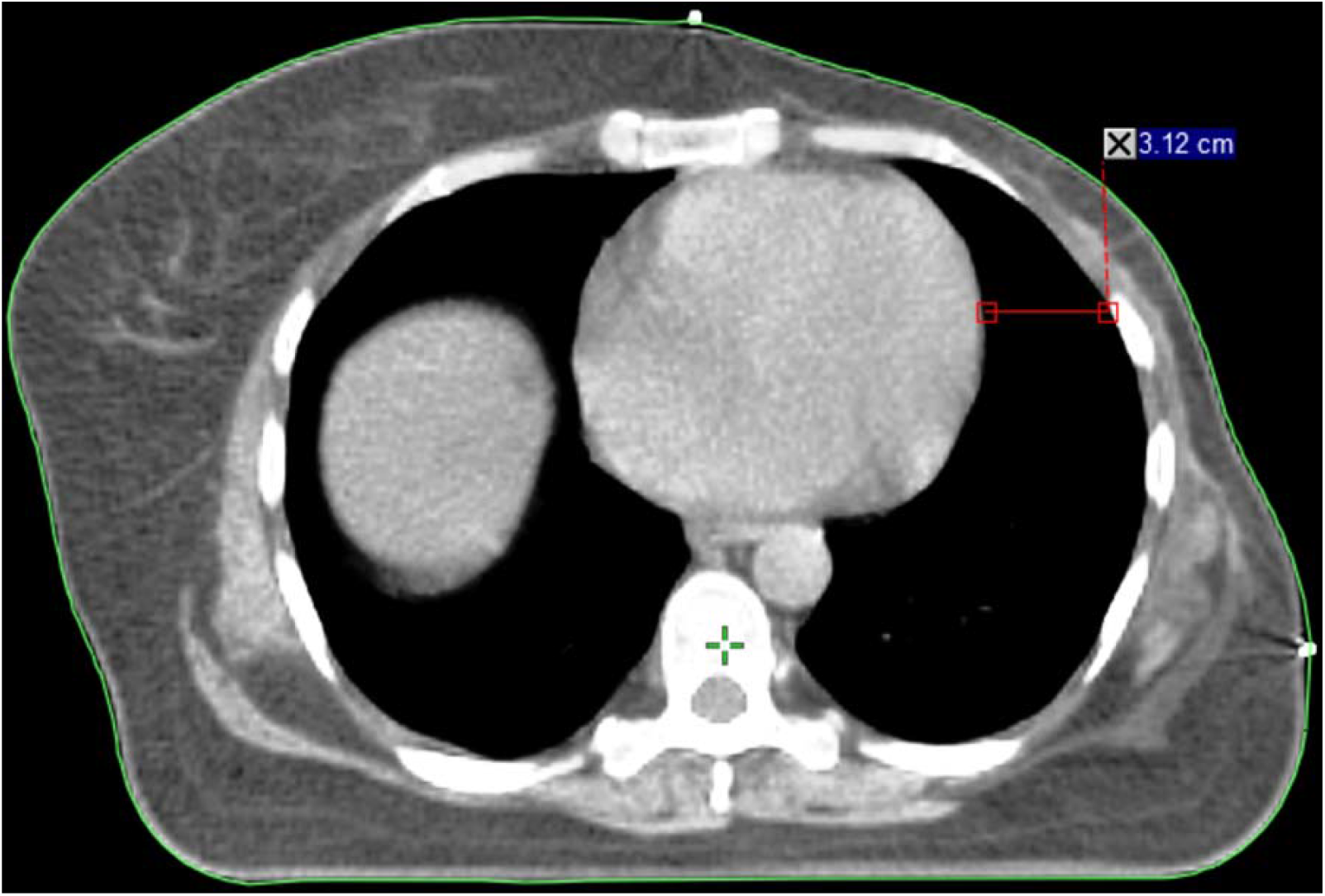
CT image acquired during free breathing (FB). This axial CT shows a reduced heart to chest wall distance of 3.12 cm, with the heart positioned closer to the left chest wall and within the high-dose region, highlighting the need for heart-sparing techniques in left-sided breast radiotherapy.

**FIGURE 3:**
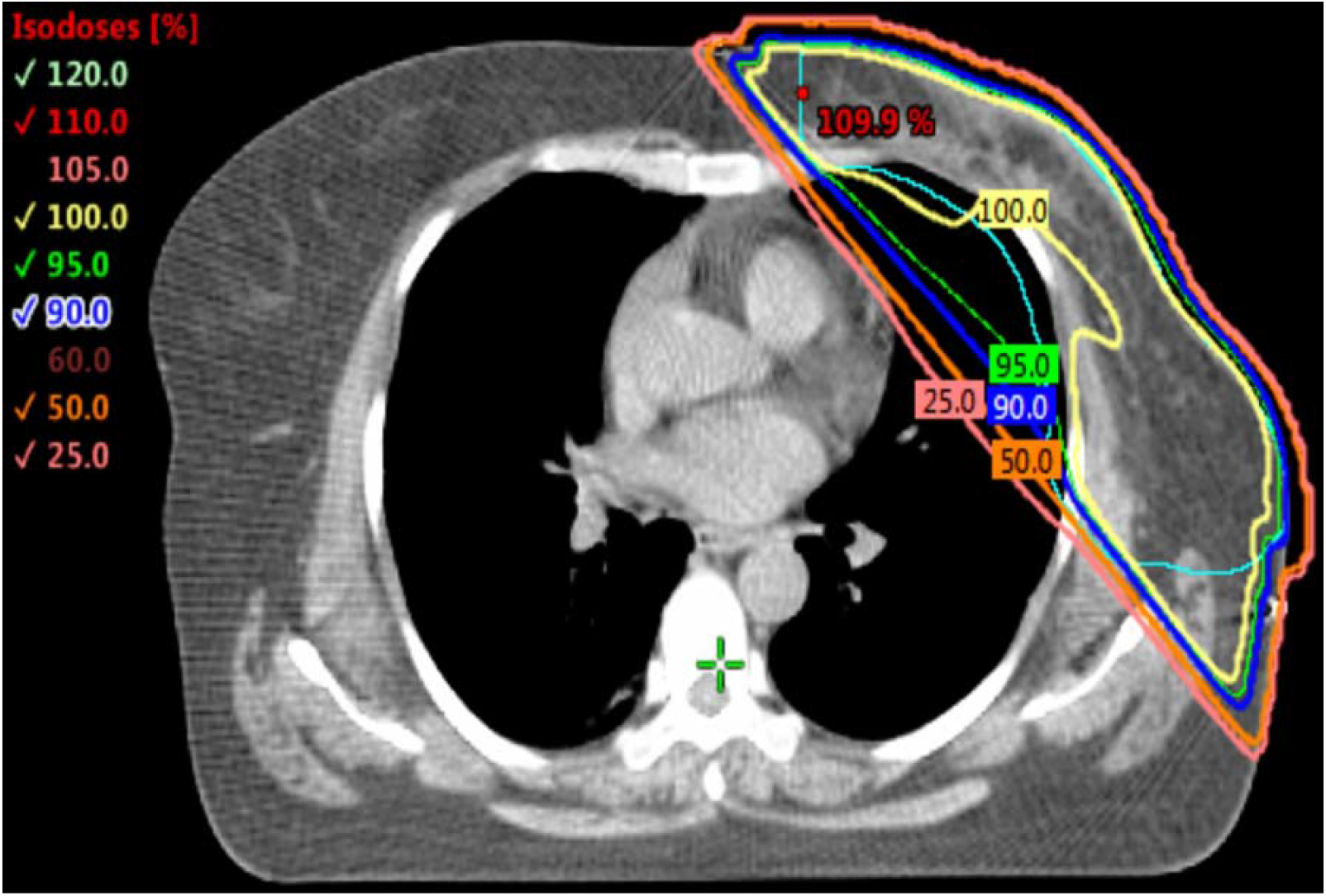
Isodose distribution from a 3D-conformal radiotherapy (3D-CRT) plan using vDIBH. The axial isodose map demonstrates homogeneous dose coverage to the planning target volume (PTV) with effective sparing of the heart and ipsilateral lung, confirming the benefit of combining 3D-CRT with vDIBH for cardiac dose reduction.

## 4. Discussion

This study demonstrates that 3D-CRT combined with vDIBH is an effective and practical technique for delivering ultra-hypofractionated whole breast radiotherapy in patients with left-sided breast cancer. The use of vDIBH allowed for robust target coverage while significantly reducing radiation exposure to the heart and ipsilateral lung. Our findings align with previous studies showing that vDIBH increases the distance between the heart and chest wall, thereby lowering cardiac doses. In comparison to the pooled average heart dose of 4.7 Gy reported in previous DIBH studies (Smyth et al., 2015), our cohort demonstrated marginally lower mean heart doses, reinforcing the effectiveness of the technique. Lung sparing was also achieved without compromising target coverage, in agreement with prior literature demonstrating reduced lung dose through thoracic expansion during deep inspiration. Long-term data from the Early Breast Cancer Trialists’ Collaborative Group (EBCTCG) showed that patients treated with radiotherapy in the 1960s and 1970s experienced a 30% increase in cardiac mortality [2]. Although modern CT-based planning techniques have reduced unnecessary cardiac exposure, studies with over 10 years of follow-up are still needed to assess the true incidence of late cardiac events [1]. Darby et al. reported that each 1 Gy increase in mean heart dose is associated with a 7.4% increase in the rate of major coronary events, including myocardial infarction and cardiac death [6,12]. Similarly, Sardaro et al. estimated a 4% increase in the risk of late cardiac disease per 1 Gy [6].

Our approach to vDIBH implementation involved a simple, low-cost setup using in-room lasers, skin tattoos, and cine mode imaging for intra-fraction verification. This ensured reproducible breath-hold positioning without requiring expensive respiratory monitoring equipment. Patient tolerance and compliance were excellent, and breath hold reproducibility was maintained throughout treatment. No increase in treatment time or workflow disruption was observed, supporting the feasibility of incorporating vDIBH into standard radiotherapy practices even in resource-limited centers.

While the heart is the primary concern in left-sided breast radiotherapy, minimizing dose to the ipsilateral lung is also critical. Our data confirmed that vDIBH also achieved effective lung sparing, consistent with prior reports. Additionally, contralateral breast dose remained low. Although the long-term impact is still debated, some evidence such as from atomic bomb survivor data suggests that doses between 1-4 Gy may modestly increase the risk of contralateral breast cancer after more than 15 years [10]. Visual coaching and feedback methods, such as video goggles, have been shown to further enhance breath hold reproducibility and treatment accuracy in left-breast cancer patients undergoing DIBH [17]. These techniques may offer additional benefits when integrated into standard workflows.

However, this study is limited by its retrospective design, relatively small sample size, and absence of a direct free-breathing comparison. Clinical outcomes such as cardiac toxicity, secondary malignancies, or cosmetic effects were not evaluated. Future prospective studies with larger cohorts and long-term follow-up are warranted to validate these dosimetric benefits and assess their clinical significance. In summary, our findings support the clinical utility of 3D-CRT with vDIBH, which offers a simple, effective, and resource efficient method for delivering hypofractionated left-sided breast radiotherapy while minimizing radiation dose to critical organs. Its ease of implementation, patient compliance, and cost-effectiveness support its broader adoption in modern clinical practice.

## 5. Conclusion

The combination of 3D-CRT with vDIBH has demonstrated clear dosimetric and clinical advantages in the treatment of left-sided breast cancer. This technique offers excellent target coverage while significantly reducing radiation exposure to the heart and lungs, which is critical for minimizing long-term cardiopulmonary risks.

Our approach utilized a cost-effective, easily implementable setup involving in-room lasers, skin tattoos, and cine mode imaging for intra-fraction verification. This ensured reproducible breath-hold positioning without the need for expensive respiratory gating systems or prolonging treatment times. The technique was well tolerated by patients and did not impact daily throughput, underscoring its feasibility in busy clinical environments. The safety, simplicity, and effectiveness of vDIBH support its routine use in left-sided breast radiotherapy, including in resource-limited settings. Broader adoption of this approach can improve treatment quality, reduce toxicity, and contribute to better long-term outcomes for breast cancer patients. However, despite the dosimetric benefits, the risk of cardiac toxicity from residual heart exposure remains an important consideration. This underscores the need for careful treatment planning and continued efforts to optimize heart sparing while maintaining target coverage.

## Data Availability

All data produced in the present study are available upon reasonable request to the authors

## Acknowledgements

none

## Funding

none

## Conflict of interest

none declared

## Ethics

The procedure followed were in accordance with the ethical standards of the responsible committee on human experimentation and the Helsinki Declaration of 1964, as revised in 2013. The study was approved by the institutional Ethics Committee of Meherbai Tata Memorial Hospital, Jamshedpur(IEC/2025/July 25), dated July 25, 2025 and waiver of consent was obtained as this was retrospective study.

## Notes

### Competing Interest Statement

The authors have declared no competing interest.

### Funding Statement

This study did not receive any funding

### Author Declarations

Ethics committee/IRB of Meherbai Tata Memorial Hospital waived ethical approval for this work

### Summary of Updates

Revised paper allignment , and some minor changes in figures

